# Preregistered analytic plan: the gut microbiome and acute kidney injury in sepsis

**DOI:** 10.1101/2024.04.04.24305205

**Authors:** Katherine M. Winner, Rishi Chanderraj, Ying He, Mark Nuppnau, Robert J. Woods, Michael Heung, Jennifer A. Schaub, Michael W. Sjoding, Robert P. Dickson

**Affiliations:** Department of Microbiology and Immunology, University of Michigan Medical School, Ann Arbor, MI, 48109, USA; Division of Pulmonary and Critical Care Medicine, Department of Internal Medicine, University of Michigan Health System, Ann Arbor, MI, 48109, USA; Medicine Service, Infectious Diseases Section, Veterans Affairs (VA) Ann Arbor Healthcare System, Ann Arbor, Michigan, USA; Division of Infectious Diseases, Department of Internal Medicine, University of Michigan Health System, Ann Arbor, MI 48109, USA; Center for Computational Medicine and Bioinformatics, University of Michigan Medical School, Ann Arbor, Michigan; Division of Nephrology, Department of Internal Medicine, University of Michigan Health System, Ann Arbor, MI, 48109, USA; Institute for Healthcare Policy and Innovation, University of Michigan, Ann Arbor, Michigan; Weil Institute for Critical Care Research & Innovation, Ann Arbor, MI, USA

## Abstract

**Overview:** We here share a pre-registered analytic plan for a matched case-control study nested in a retrospective cohort of hospitalized patients with suspected sepsis. We will compare gut microbiota (measured near the time of admission) among patients with sepsis who do and do not develop sepsis-induced acute kidney injury.

**Rationale:** Sepsis afflicts nearly 50 million people annually, and sepsis-induced acute kidney injury (AKI) is a frequent complication that contributes to morbidity, mortality, and increased healthcare costs. Despite the clinical significance of AKI in sepsis, we still do not understand why some patients with sepsis develop AKI while others do not. The gut microbiome has been implicated in other clinical features and sequelae of sepsis, but to date has not been studied in sepsis-induced AKI.

**Objective:** To determine whether gut microbiota predict AKI in patients with suspected sepsis.

**Hypothesis:** We hypothesize that among patients with suspected sepsis, gut bacterial density and identity (at the time of admission) predict the onset and severity of AKI.

**Study design:** We will perform a matched case-control study nested in an observational cohort. The cohort includes patients admitted to the University of Michigan in 2016-2020 with suspected sepsis. We will divide patients into cohorts that did and did not develop AKI. We will derive matched cohorts based on relevant clinical covariates. We will characterize their gut microbiota using 16S rRNA gene amplicon sequencing of rectal swabs obtained within 24 hours of AKI onset. We will compare admission gut microbiota across these matched cohorts to test our primary hypothesis.

## Body

### Overview

We have designed a matched case-control study nested in a retrospective cohort of hospitalized patients with suspected sepsis. We will test the hypothesis that gut microbiota (measured near the time of admission) predict the onset and severity of acute kidney injury (AKI) among patients with sepsis.

### Rationale

Sepsis afflicts nearly 50 million people annually, and AKI is a frequent complication that contributes to morbidity, mortality, and increased healthcare costs^1–4^. Despite the clinical significance of AKI in sepsis, we still do not understand why some patients with sepsis develop AKI while others do not^5^, limiting our ability to predict and prevent its onset and outcomes.

The gut microbiome represents a tremendous source of biological variation among hospitalized patients. In both animal models^6^ and observational human studies^7^, gut microbiota are correlated with the severity of systemic injury and risk of death in critical illness. Our research group has recently demonstrated that gut microbiota predict the severity of response (weight loss, temperature) in a murine model of sepsis, confirming the relationship between gut microbiota and heterogeneity in sepsis pathogenesis^8^.

Relevant to the pathogenesis of sepsis-associated AKI (SA-AKI), the gut microbiome: 1) participates in the calibration of systemic immunity^9^, 2) is a metabolic “organ” that generates hundreds of systemically active metabolites^10,11^, 3) is a well-recognized source of uremic toxins^12^, 4) is a source of systemically translocated bacteria and bacterial products in sepsis^13^, and 5) provides colonization resistance against secondary (nosocomial) infections^14^. Several studies have demonstrated that gut microbiota participate in the pathogenesis of AKI caused by conditions other than sepsis (e.g., ischemia-reperfusion injury)^15–19^. Yet, the contribution of gut microbiota to the pathogenesis of SA-AKI has been unexplored to date, both in experimental models and in human cohort studies.

### Objective

To determine whether gut microbiota predict AKI in patients with suspected sepsis.

### Hypothesis

We hypothesize that among patients with suspected sepsis, gut bacterial density and identity (at the time of admission) predict the onset and severity of AKI.

### Study design (overview)

We will perform a matched case-control study nested in an observational cohort. The cohort includes patients admitted to the University of Michigan in 2016-2020 with suspected sepsis. We will divide patients into cohorts that did and did not develop AKI. We will derive matched cohorts based on relevant clinical covariates. We will characterize their gut microbiota using 16S rRNA gene amplicon sequencing of rectal swabs obtained within 24 hours of AKI onset. We will compare admission gut microbiota across these matched cohorts to test our primary hypothesis.

### Access to clinical data

To identify our cohort and perform matching, we used a structured language query of the University of Michigan’s Research Data Warehouse. We collected the following clinical information for patients meeting criteria for suspected sepsis (defined below) in 2016-2020: demographics, past medical history, laboratory values, vital signs from nursing flowsheets, medical administration records, and admission metadata.

### Specimen acquisition

Biospecimens in this study are rectal swabs performed as a routine aspect of clinical care. These swabs were performed routinely for surveillance of Vancomycin-Resistant *Enterococcus* (VRE) in patients admitted to any of seven high-risk inpatient units (Critical Care Medical Unit, Surgical Intensive Care Unit, Neurologic Intensive Care Unit, Coronary Care Unit, Trauma-Burn Intensive Care Unit, Cardiothoracic Surgery Unit, Intensive Care Step-down Unit), bone marrow transplant ward, or oncology ward. Rectal swab specimens were collected on admission by nursing staff with the BD™ ESwab Regular Collection Kit (Franklin, NJ). The prescribed practice at our institution during the study period was to acquire rectal swabs from patients in left lateral decubitus position. A rectal swab was inserted through the rectal sphincter 2-3 cm, rotated 360°, withdrawn and checked for the presence of fecal soilage. Following testing for VRE by the University of Michigan Clinical Microbiology Laboratory, the contents of the swabs are dislodged into a buffer solution with glycerol and stored at -80°C. These banked rectal swabs have been previously used for human microbiome studies^8,20–22^.

### Study population

The study population consists of adult subjects admitted to the University of Michigan in 2016-2020 with clinically suspected sepsis and a rectal swab collected close to admission available for analysis in a banked repository. We defined suspected sepsis based on a modification of the Rhee criteria^23^: patients were identified as having suspected sepsis if blood cultures were collected within 24 hours of admission and four consecutive days of antibiotic treatment was administered within ±2 days of the day of blood culture collection. We excluded organ failure from the Rhee criteria to ensure a wide range of disease severity and include a mix of subjects with and without organ failure. To ensure our analysis is representative of the gut microbiome around the time of AKI onset, we restricted our cohort to patients whose swab was collected no more than 24 hours after AKI onset.

Because we are specifically interested in *de novo* AKI, we excluded patients with end-stage renal disease (ESRD) and chronic kidney disease (CKD). Because in many instances we could not determine if a patient had ESRD or CKD prior to admission, we also excluded patients with abnormal creatinine (>1.2 mg/dL) throughout their entire hospital stay, recognizing that this is a conservative threshold and will exclude some patients with non-resolving AKI. We also excluded patients if they were transferred from an outside institution, because 1) we lacked sufficient medical history for these patients (pertinent for the identification of covariates such as medical comorbidities) and 2) we could not be confident regarding timing of sepsis onset, AKI onset, and antibiotic exposures. Finally, we excluded the fourth or greater hospital readmission and any readmissions that were fewer than 90 days apart.

### Definition of AKI

We defined and staged AKI using the Kidney Disease Improving Global Outcomes (KDIGO) consortium definition of AKI, which defines AKI onset as a ≥ 50% increase in baseline serum creatinine within seven days of admission^24^. The KDIGO definition of AKI builds upon prior RIFLE^25^ and AKIN^26^ criteria for defining and staging AKI and has been used successfully in numerous studies. The National Kidney Foundation-Kidney Disease Outcomes Quality Initiative (NKF-KDOQI) supports the use of the KDIGO AKI guideline definition and staging strategy for epidemiologic comparisons across populations and over time^27^.

When determining how to define baseline creatinine for AKI staging, we considered three options: 1) outpatient creatinine measured prior to hospitalization, 2) minimum creatinine measured within 7 days prior (“rolling window”), and 3) absolute minimum creatinine measured during hospitalization. Although using outpatient creatinine measurements would be a rigorous approach, nearly 30% of patients in the cohort did not have outpatient creatinine information available, limiting our ability to use this definition. Using the “rolling window” approach fails to capture patients who already had developed AKI by the time of their presentation, while the absolute minimum creatinine approach is able to capture patients with resolving AKI. When modeling the “rolling window” approach, we observed that numerous patients with AKI were misclassified as not having AKI compared to patients as defined by the absolute minimum approach. We thus decided to define baseline creatinine as the patient’s absolute minimum creatinine throughout their hospitalization, recognizing that an inherent limitation of this approach is potential omission of patients who present with AKI that does not resolve during hospitalization. While we considered a “hierarchical” approach in which outpatient creatinine is used when available and minimum inpatient creatinine is used in all other patients, we elected to use a uniform definition for the sake of standardization and minimization of bias across the entire cohort.

### Identification of cohort

From the initial 17,952 patients, 1,458 met inclusion criteria (**Figure 1**). We defined cases as patients who met KDIGO criteria for stage 2 or 3 AKI (≥ 100% or ≥ 200% increase in baseline creatinine within 7 days, respectively) and controls as patients that did not develop AKI. We chose to exclude stage 1 AKI in our definition of both AKI cases and controls, because stage 2 and 3 AKI are associated with relatively higher risk of major adverse kidney events and death^28,29^, and we sought to ensure sufficient clinical separation between cases and controls. We used coarsened exact matching to match cases and controls 1:2 by age (age was discretized and binned into tertiles), gender, and unit of admission. We chose to match on age and gender because these are predictors of AKI^30,31^ and unit of admission as a partial proxy for admission diagnosis and provider practice. We did not match on antibiotic treatment because antibiotics directly alter gut microbiota; thus matching on this exposure would obscure the very source of variation we intend to study. We chose not to match on baseline kidney function (eGFR) because 1) the calculated eGFR is based on variables already incorporated into our matching strategy (age, sex, creatinine)^32^ and 2) cases and controls are already similar in eGFR (**Table 1**). From the cohort, 435 patients were matched (145 cases, 290 controls). The matched variables, baseline kidney function (eGFR), and burden of medical comorbidities are evenly distributed among cases and controls (**Table 1**). This balance of baseline patient demographics and underlying kidney function demonstrated an equivalent susceptibility to AKI between cases and controls.

**Table 1.** Cohort demographics

| **Table 1 Cohort demographics** |  |  |  |
| --- | --- | --- | --- |
|  | **No AKI** | **AKI** | **p Value** |
| *n* | 290 | 145 |  |
| Male | 150 (51.7) | 75 (51.7) | 1 |
| Age, years | 55.62±16.74 | 56.12±16.69 | 0.767 |
| Unit |  |  | 1 |
| BICU | 2 (0.7) | 1 (0.7) |  |
| BMT | 18 (6.2) | 9 (6.2) |  |
| CCU | 20 (6.9) | 10 (6.9) |  |
| CVC | 14 (4.8) | 7 (4.8) |  |
| MICU | 72 (24.8) | 36 (24.8) |  |
| ONC | 96 (33.1) | 48 (33.1) |  |
| SICU | 44 (15.2) | 22 (15.2) |  |
| STEP_DOWN | 24 (8.3) | 12 (8.3) |  |
| Non-Caucasian | 52 (17.9) | 21 (14.6) | 0.458 |
| eGFR* | 94.19±21.43 | 93.21±32.91 | 0.711 |
| APACHE IV | 45.87±21.27 | 70.74±26.38 | <0.001 |
| Hospital Death | 26 (9.0) | 18 (12.4) | 0.339 |
| Charlson Index | 3.40±3.16 | 3.20±3.27 | 0.539 |
| Myocardial Infarction | 22 (7.6) | 17 (11.7) | 0.213 |
| Congestive Heart Failure | 43 (14.8) | 22 (15.2) | 1 |
| Peripheral Vascular Disease | 11 (3.8) | 16 (11.0) | 0.006 |
| Chronic Pulmonary Disease | 71 (24.5) | 33 (22.8) | 0.781 |
| Mild Liver Disease | 26 (9.0) | 21 (14.5) | 0.113 |
| Moderate-severe Liver Disease | 13 (4.5) | 12 (8.3) | 0.166 |
| Diabetes | 54 (18.6) | 39 (26.9) | 0.063 |

\*eGFR was calculated with the CKD-EPI creatinine equation (2021) Data are presented as n, mean±SD or n (%). APACHE: Acute Physiology and Chronic Health Evaluation

**Figure 1.**
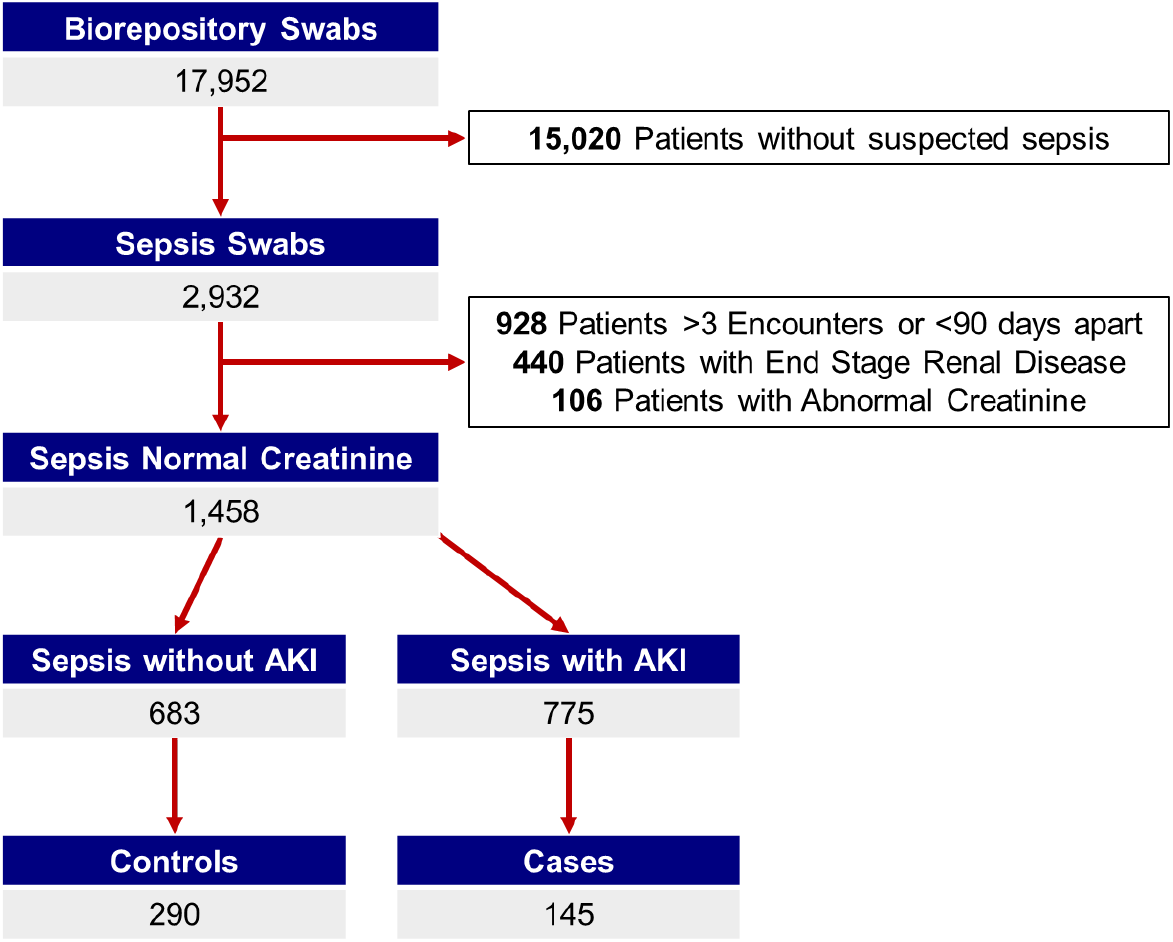
Cohort Overview. We identified a cohort of 1,458 patients with suspected sepsis admitted to the University of Michigan in the years 2016-2020. We excluded the fourth or greater admission and any readmissions less than 90 days apart. We also excluded patients with known end-stage renal disease or an abnormal creatinine (>1.2 mg/dL) throughout their hospitalization.

### Sample size considerations

We included all available cases and two matched controls from our cohort. Study sample size was thus governed by rectal swab collection and AKI incidence. We used a 1:2 (case:control) matching strategy to improve statistical power. To confirm the plausibility of our detecting a significant difference in gut microbiome composition between patients who develop AKI and those who do not, we performed a formal power calculation. Based on preliminary data generated from 116 swabs from this same repository^8,20–22^, relative abundance of *Lachnospiraceae* (a prominent bacterial family associated with sepsis severity in animals and humans) was 6.5% ± 7.4%^8^. Thus, assuming an alpha of 0.05, we anticipate the following minimum detectable difference according to various estimated power thresholds:

| **Estimated Power** | **Minimum Detectable Difference (%)** |
| --- | --- |
| 80% | 2.1% |
| 90% | 2.4% |
| 95% | 2.7% |
| 99% | 3.2% |

Alternately, we consider the Shannon Diversity Index (an integrative index of community richness and evenness). In the same pilot study of 116 rectal swabs from a similar human cohort, the mean Shannon Diversity Index was 2.7 ± 0.66^8^. We thus anticipate the following minimum detectable differences for Shannon Diversity Index:

| Estimated Power | Minimum Detectable Difference (%) |
| --- | --- |
| 80% | 0.188 |
| 90% | 0.218 |
| 95% | 0.243 |
| 99% | 0.288 |

We thus anticipate we will have adequate statistical power for our primary comparisons.

## Generation of microbiome data

### Bacterial DNA Isolation

We will extract genomic DNA from rectal swabs resuspended in 360 μL ATL buffer (Qiagen DNeasy blood and tissue kit) and homogenized in fecal DNA bead tubes. We will include multiple negative control specimens (unused swabs, isolation controls, AE buffer, sterile water, blank wells) to identify potential procedural and sequencing contamination. We will use the ZymoBIOMICS Microbial Community DNA Standard (Zymo Research) as a positive control. We anticipate having to use multiple extraction kits, we will use the fewest possible and keep track of which kits were used to determine any potential batch effects.

### Bacterial Density Quantification

We will quantify extracted bacterial DNA from rectal swabs using a QX200 Droplet Digital PCR System (BioRad, Hercules, CA) as previously described^20,21^. Primers and cycling conditions will be performed according to a previously published protocol^33^. To summarize, we will use primers 5′-GCAGGCCTAACACATGCAAGTC-3′ (63F) and 5′-CTGCTGCCTCCCGTAGGAGT-3′ (355R). The cycling protocol is as follows: 1 cycle at 95°C for 5 min, 40 cycles at 95°C for 15 s, and 60°C for 1 min, 1 cycle at 4°C for 5 min, 1 cycle at 90°C for 5 min, all at a ramp rate of 2°C/s. We will use the BioRad C1000 Touch Thermal Cycler for PCR cycling and detect droplets using the automated droplet reader (BioRad, catalog no.1864003), using Quantasoft™ Analysis Pro (version 1.0.596) for quantification. Both sterile water controls, as well as isolation controls will be run alongside rectal swab specimens.

### 16s rRNA gene sequencing

We will amplify the V4 region of the 16S rRNA gene from extracted genomic DNA using published primers and the dual-indexing sequencing strategy as described previously^34^. We will use the Illumina MiSeq platform (San Diego, CA) with a MiSeq Reagent Kit V2 (500 cycles), according to the manufacturer’s instructions with modifications found in the standard operating procedure of the laboratory of Patrick Schloss^34,35^. Given the cohort size, samples will be sequenced over multiple sequencing runs. To account for batch effect, we will randomize samples across runs (to avoid false clustering, e.g. false clustering of sequentially collected specimens) and include the ZymoBIOMICS Microbial Community DNA Standard (positive control) and negative controls to assess run-to-run variation. We will retain the run number as a variable in our analysis and use this to compare α-and β-diversity between runs to assess batch effects. If batch effects are large, we will use run number as a variable in our multivariate logistic regression analyses.

### 16S gene amplicon analysis

We will process and analyze sequence data using the software *mothur*^1^ according to the standard operating procedure for MiSeq sequence data^2,3^. We will follow the *mothur* standard operating procedure without deviation. To summarize, we will use the VSEARCH^4^ algorithm to detect chimeras using abundant sequences as our reference. We will remove sequences flagged as chimeric from all samples. We will use the SILVA rRNA database^5^ as a reference for sequencing alignment and perform pairwise alignment with the Needleman-Wunsch^6^ algorithm. We will pass a distance matrix to the OptiCLUST clustering algorithm^7^ with a 97% distance threshold to cluster sequences into “operational taxonomic units” (OTUs). We will classify OTUs using the *mothur* implementation of the Ribosomal Database Project (RDP) classifier and RDP taxonomy training set 18 (trainset18_062020.rdp.fasta, trainset18_062020.rdp.tax), available on the *mothur* website.

After clustering and classification of sequencing data, will perform all analyses in R. We will evaluate differences in community structure with permutational multivariate analysis of variance (PERMANOVA) using the *vegan* package in R^45^. We will also perform resampling of multiple generalized linear models with the *mvabund*^46^ package in R to look for individual OTU differences between communities. We will compare the community structure of negative controls to rectal swab specimens and confirm statistically significant differences using orthogonal techniques of hypothesis testing (PERMANOVA and *mvabund*). If we find that negative-control specimens are dominated by a single OTU (indicating the presence of true contamination rather than stochastic sequencing “noise”^47^), we will perform our primary analyses with and without exclusion of that OTU as a sensitivity test to determine the influence of contamination on our findings.

We will not reflexively perform a “background subtraction” step, as is sometimes performed in low-biomass microbiome studies. Our rationale is that 1) many 16S-classified taxa present in negative control specimens may also represent “true” taxa present in lower gut specimens, 2) well-to-well contamination is a known phenomenon on Illumina sequencers, thus introducing “true” signal into negative control specimens^48^, 3) given the compositional nature of microbiome data, removal of any OTU directly results in the “inflation” of all other taxa, including possible contaminants not detected in negative control specimens, and 4) the presence of background taxa in specimens may be an indirect proxy for *low bacterial density*, which is itself a biologically and clinically significant feature of microbial communities^20^. For these reasons, we will instead prioritize transparency, reporting all taxa detected in both specimens and negative controls. We will make our sequencing data (and all relevant metadata) publicly available prior to submission of our first manuscript so that reviewers and readers can directly compare taxa detected in our specimens and negative controls. We will use absolute 16S rRNA gene quantification (using ddPCR as above) as an additional, orthogonal measurement to discriminate “true” and “false” microbial signal in our specimens.

## Primary hypothesis

In hospitalized patients with sepsis, the community composition of gut microbiota differs among those who develop AKI as compared to those who do not.

### Analytic approach

We will use PERMANOVA to compare community composition of cases and controls, stratified by matched pair. We will use the Bray-Curtis dissimilarity index as our measure of compositional dissimilarity. We will perform our analysis at both the OTU and family level of taxonomic classification. We will use 10,000 permutations for each execution of PERMANOVA. We will interpret a P value of < 0.05 as indicative of a significant difference in community composition.

Should PERMANOVA reveal no difference in community composition across cases and controls, we will interpret this as falsification of our primary hypothesis (i.e., as evidence that within our cohort, the community composition of gut microbiota *does not differ* among septic patients that do and do not develop AKI). Wherase should PERMANOVA analysis identify a global difference in gut community composition between case and control subjects, we will determine the family-level differences in community composition that explain this difference using a random forest classification model built with the *ranger* package^49^. We will run the random forest model 100 times using Mean Decrease in Accuracy as our index of feature importance. We will construct two models using this method: one using relative abundance of bacterial taxa and the other using absolute abundance (relative ^*^ bacterial density) of bacterial taxa. We will correct for feature importance bias and assign significance values to key bacterial taxa with a permutation importance heuristic (PIMP)^50^ and identify bacterial taxa that meet a threshold of significance p<0.01.

We will then use a multivariable conditional logistic regression model (conditioned on matched pair) to determine if the relative and absolute abundance characteristics are *independently* associated with the onset of AKI. We will include the abundance of the bacterial families that meet a threshold significance of p<0.01 identified by random forest and the following covariates: APACHE IV score, Charlson comorbidity index, and the administration of known nephrotoxins and vasopressors.

We will also test the relative abundance of key (pre-specified) bacterial taxa across cases and controls. In pilot studies of a murine model of sepsis, we identified *Lachnospiraceae, Ruminococcaceae, Lactobacillaceae, Enterobacteriaceae*, and *Clostridiaceae* as bacterial families associated with sepsis severity. We will use the Wilcoxon signed-rank test to compare both relative and absolute abundance of the above bacterial families, as well as those that meet a threshold significance of p<0.01 identified by random forest, between cases and controls. Given the exploratory nature of this analysis, we will adjust for multiple comparisons using the Bonferroni correction.

## Secondary hypotheses

1. In hospitalized patients with sepsis, the *density* of admission gut microbiota is higher among those who develop AKI as compared to those who do not. <u>Approach:</u> Our lab has demonstrated that the bacterial density of rectal swabs is highly variable, and that this variability is of clinical significance^20^. We will use the log-transformed number of 16S copies per rectal swab (quantified using ddPCR as above) to determine the bacterial density of admission rectal swabs. We will compare the bacterial density between case and control subjects using paired t-testing to determine if there is a significant difference between the groups. We will interpret a P value of <0.05 as indicative of a significant difference in bacterial density.
2. In hospitalized patients with sepsis, the *diversity* of admission gut microbiota is lower among those who develop AKI as compared to those who do not. <u>Approach:</u> In a systematic review of 69 studies on gut microbiota composition in patients with chronic kidney disease, more than half of the studies reported that gut diversity is significantly decreased in CKD (from early to advanced stages) compared to healthy individuals^51^. We will thus calculate the Shannon Diversity Index (a measure of both evenness and richness), the Chao1 Index (a measure of richness), and the dominance (a measure of evenness) of admission rectal swabs using the *vegan* package in R to establish whether the evenness and richness of the gut microbiome is associated with AKI onset. We will compare these three complementary indices between case and control subjects using paired t-testing and interpret a P value of <0.05 as indicative of a significant difference in bacterial diversity.
3. Differences in *both* density and diversity of baseline gut microbiota contribute to the onset of AKI in septic patients. <u>Approach:</u> We know that variability in gut bacterial density and diversity is clinically significant^20,51^, but we do not know which of these features of the microbiome are more influential in determining AKI onset. Using a multivariable *conditional* logistic regression model (conditioned on matched pair) we will determine if both bacterial density and diversity are independently associated with the onset of AKI. The multivariable logistic regression model will include a primary outcome of AKI with the covariates APACHE IV score, Charlson comorbidity Index, administration of known nephrotoxins and vasopressors, gut bacterial density, and gut bacterial diversity. Including gut bacterial density and diversity in the logistic regression will inform us which feature of the microbiome matters more for the onset of AKI in septic patients.

## Exploratory analyses

This project will generate the largest dataset to date for hospitalized patients with suspected sepsis that contains both gut microbiome data and patient-matched clinical exposure and outcome data. It will thus be a robust source of data for use in hypothesis-generating analyses. In exploratory analyses, we will compare the relationship between antibiotic exposure, gut microbiota, and important outcomes: mortality, ventilator-free days, and organ failure-free days. All correlations identified via these exploratory (non-AKI) analyses will be considered provisional until validated in independent cohorts, animal models, and other subsequent investigations.

## Data Availability

All data produced in the present work are contained in the manuscript.

## Notes

### Competing Interest Statement

The authors have declared no competing interest.

### Funding Statement

This study did not receive any funding.

### Author Declarations

The University of Michigan Institutional Review Board has approved this study (HUM00222404). The same review body deemed the study exempt from needing consent under 45 CFR section 46, category 4 (secondary use of identifiable data).

